# Triage with AI: A Rule-out Framework Quantifying the Risks and Benefits of Screening Mammogram Automation

**DOI:** 10.1101/2025.04.25.25326396

**Authors:** Micheal H. Bernstein, Maggie Chung, Adam Yala, Grayson L. Baird

## Abstract

**Background:** AI has been proposed as a triage or “rule out” device to reduce radiologist workload, but it is presently unclear how an AI triage threshold should be determined. We present a framework for determining an optimal threshold.

**Materials and Methods:** 114,229 bilateral 2D digital screening mammograms were retrospectively analyzed from 2006-2023. All mammograms were given an AI score using Mirai, an open-source deep-learning model. Several metrics were examined using two thresholds for determining ruled out versus retained cases: 1) Caseload Reduce Rate (CRR; percent of caseload reduced due to rule-out), 2) Gross AI False Omission Rate (G-FOR; probability of a patient having breast cancer if ruled out), 3) AI Net False Omission Rate (N-FOR; probability of a patient having breast cancer if ruled out and the radiologist would have caught in standard care [i.e. no triage].), 4) AI Adjusted Net False Omission Rate (30%) (AN-FOR[30%]; N-FOR adjusted for the hypothetical scenario where radiologists detect an extra 30% of breast cancers among AI retained cases). The two thresholds were severity scores of 0.2 (Yuden’s J) and 0.05 (AN-FOR[30%]=0). The former is mathematically optimal; the latter reflects a threshold where AI triage does not introduce any total increase in False Negatives.

**Results:** At the 0.20 threshold, G-FOR, N-FOR, and AN-FOR(30%) were 0.26%, 0.017%, and 0.14%, respectively (223, 141, and 121, respectively, missed cancer cases) and CRR=75%. At the 0.05 threshold, the G-FOR, N-FOR, and AN-FOR (30%) are 0.12%, 0.07%, and 0.00% (49, 30, and 0, respectively, missed cancer cases) and CRR=36%.

**Conclusion:** We demonstrate how radiology practices can consider the trade-offs of using different AI scores triage thresholds. At the AN-FOR rate of 30%, the Yuden’s J threshold results in 121 additional missed cancers for a 75% caseload reduction. We estimate no additional missed cancers at a 36% caseload reduction.

## Introduction

The increasing use of artificial intelligence (AI) in radiology has prompted considerations about its potential in addressing the field’s mounting challenges. In recent years, the workload of radiologists has grown significantly. For instance, one study found that the workload for on-call radiologists in the Emergency Department quadrupled between 2006 and 2020.^1^ Another study^2^ that examined billed work relative value units (RVUs) among more than 35,000 academic radiologists found a 60% increase in workload from 2008 to 2020. This growing burden contributes to rising rates of burnout.^3,4^ Furthermore, the number of individuals entering radiology residency has not kept pace with the rise of imaging volume,^5^ creating a growing workforce imbalance that is unlikely to be resolved in the near future.

AI has the potential to alleviate some of this burden. Studies have found that AI can reduce medical imaging interpretation times.^6–8^ One promising application to improve efficiency is using AI as a triage or “rule out” device. By identifying mammograms that are extremely low-risk, AI can reduce the number of cases that require interpretation by a radiologist. Low-risk, non-triaged cases can be safely ruled out and recorded as negative (i.e., no evidence of abnormality) without radiologist review. This "rule-out" approach is well-suited for pathologies with a low prevalence rate, where there are many true negative cases that can be ruled out at the cost of very few false negatives. Thus, AI triage may be suited for screening mammograms where fewer than 1% of cases are positives.^9,10^ The goal is to safely exclude the majority of normal cases and allow radiologists to concentrate on more suspicious exams.

Using AI triage for “rule out” in radiology has been proposed by several groups.^11–18^ Recently, empirical studies have suggested that AI triage can perform comparably to, and in some cases better than, standard of care where radiologists interpret all mammograms.^12,19–21^

One critical component of AI triage is determining the appropriate **Triage Threshold.** Most AI algorithms generate a continuous risk score for each image, with higher scores indicating a greater likelihood of pathology. However, where the precise cut-off should be placed for distinguishing which cases are ruled out (i.e., non-triage) versus reviewed by a radiologist (i.e., triage) remains an open question.^22–25^ Setting the threshold depends on balancing a variety of benefits and risks, which we discuss below. For clarity, these are divided into “ruled out” and “retained” cases.

In this article, we present a framework for determining the optimal AI triage threshold for screening mammogram automation. We outline key metrics to evaluate the trade-offs between benefits and risks at different thresholds. Specifically, using AI risk scores from the Mirai model applied to 114,229 screening mammograms, we simulate triage thresholds to quantify their effects on caseload reduction and cancer detection. We propose approaches for identifying the optimal threshold based on these metrics.

## Methods

University of California, San Francisco (UCSF) Institutional Review Board gave ethical approval for this Health Insurance Portability and Accountability Act–compliant study and waived the requirement for written informed consent.

### Operational definitions

*Benefits and Risks in Ruled-out Cases.* The primary benefit of ruling out cases is caseload reduction, which can be quantified as the **Caseload Reduction Rate (CRR) (Table 1)**. The higher the threshold, the higher the CRR; that is, the caseload reduction for radiologists will be higher when a more stringent (i.e. higher) threshold is set, ruling out a larger pool of cases. However, the benefit of a higher CRR must be weighed against a variety of other considerations.

**Table 1.** Key Metrics and Definitions.

| Metric | Definition |
| --- | --- |
| **Caseload Reduction Rate (CRR)** | The percentage of screening mammograms that would have been read by a radiologist under standard care but were excluded from review due to AI-based triage. |
| **AI Negative Predictive Value (AI-NPV)** | Probability of a patient not having breast cancer if ruled out by AI. |
| **AI Positive Predictive Value (AI-PPV)** | Probability of a patient having breast cancer, given that the case was retained for radiologist review. |
| **AI False Discovery Rate (AI-FDR)** | Probability of a patient not having breast cancer, given that the case was retained for radiologist review. |
| **AI Gross False Omission Rate (G-FOR)** | Probability of a patient having breast cancer if ruled out by AI. |
| **AI Net False Omission Rate (N-FOR)** | Probability of a patient having breast cancer if ruled out by AI, excluding cases that would have also been missed by radiologists without triage. |
| **AI Adjusted Net False Omission Rate (AN-FOR)** | Probability of a patient having breast cancer if ruled out, excluding cases that would have also been missed by radiologists, adjusted for breast cancer cases that AI retained but radiologists would have missed without triage. |

First, one must consider how accurate an AI is at correctly ruling out cancer, given the cancer prevalence in the population; this is reflected by the **AI Negative Predictive Value (AI-NPV)**. AI-NPV is the probability that a patient ruled out by AI truly does not have breast cancer. Some negative cases ruled out by AI might have otherwise been recalled by the radiologist in standard care (interpretating mammograms without AI triaging), potentially leading to unnecessary, costly, and stress-inducing diagnostic imaging and biopsies that turn out to be benign.

The aforementioned benefit must be carefully weighed against the **AI Gross False Omission Rate (G-FOR, or 1-AI-NPV),** which is the probability that a patient ruled out by AI actually has breast cancer. As the threshold is raised to exclude more cases, the G-FOR increases. That is, the CRR and the G-FOR come at a clear tradeoff; the more cases AI rules out, the higher the G-FOR will be. Nonetheless, it is important to note that not all cancers missed by AI in the ruled-out cases would have been detected by radiologists under **standard practice (SP**) (i.e., radiologist workflow absent an AI triaging model). That is, some cancer cases would likely have been missed regardless of whether AI triaging was used. To account for this, we define the **AI Net False Omission Rate (N-FOR)** as the G-FOR minus cancer cases that would have been missed by AI and radiologists (i.e. “deduct” cancer cases mutually missed by both radiologists and AI triage from the numerator).

### Benefits and Risks in Retained Cases

The **AI Positive Predictive Value (AI-PPV)** reflects the probability that a patient has breast cancer given that the case was retained for radiologist review. The **AI False Discovery Rate (AI-FDR, or 1-AI-PPV)** refers to the probability that a patient retained by AI for radiologist review does not actually have breast cancer. The higher the triage threshold, the more cases AI will rule out (i.e. the larger the CCR). This means that remaining (i.e. retained) cases are more likely to be true positives, which increases the AI-PPV and reduces the AI-FDR. However, decreasing the number of retained cases (and by definition also increasing the number of ruled-out cases) can have important implications for how they are interpreted.

Radiologist performance may improve as the retained reading pool size decreases due to reading fewer cases,^26^ reading an enriched batch with higher prevalence,^27–29^ and by consciously or unconsciously knowing that the cases were triaged^25^ (i.e., anchoring or automation bias). These additional cancer detections could potentially offset a portion of the cancers missed among ruled-out cases due to the use of AI triage (N-FOR). Taking this into account, the **Adjusted Net False Omission Rate (AN-FOR)** refers to the probability of a patient having breast cancer that would have been detected by a radiologist in standard practice if ruled out, adjusted for the additional cancer detections due to AI triage that would have missed in standard practice (i.e., “credit” cancer cases that radiologists would have otherwise missed without AI triaging them).

Another risk worth considering is that although radiologists are more likely to catch cancer cases they would have otherwise missed had AI not retained them, it is also likely that for the same reason, radiologists may also increase unnecessary recalls (i.e., radiologists recall non-cancer cases they would not have otherwise recalled had they not been retained by triage). ^25^

### Simulation Methods

To illustrate the trade-offs associated with different AI triage thresholds, we conducted a “simulation” using risk scores from a deep learning model with screening mammography.

#### Study Sample

We conducted a single institution retrospective review of 114,229 bilateral 2D digital screening mammograms acquired between January 2006 and January 2023. Exams with histopathologically confirmed breast cancer within 12 months of the screening mammogram were considered positive. Exams with at least 12 months of follow-up without a breast cancer diagnosis were considered negative. Based on these criteria, 864 cases (0.76%) were identified as positive.

#### AI Model

Mammograms were assessed using Mirai, an open-source deep-learning model trained to predict breast cancer risk from mammograms.^30,31^ One-year risk scores (henceforth “scores” or “Mirai scores”) were used to simulate triage thresholds.

Threshold Simulation Framework. We simulated various triage thresholds based on Mirai scores. For each threshold, we calculated the following metrics:

Caseload reduction rate (**CRR**): the percentage of screening mammograms that would have been read by a radiologist under standard care but were excluded from review due to AI-based triage. CRR=Total cases ruled out /Total cases.

AI Negative predictive value (**AI-NPV**): probability of a patient not having breast cancer if ruled out. AI-NPV = TN / (FN + TN).

AI Positive predictive value (**AI-PPV**): probability of a patient having breast cancer given if retained. PPV = TP / (TP + FP).

Gross AI False omission rate (**G-FOR**): probability of a patient having breast cancer if ruled out. G-FOR = (1-NPV) = FN / (FN + TN).

AI Net False omission rate (**N-FOR**): probability of a patient having breast cancer if ruled out and the radiologist would have caught it.

AI Adjusted Net False omission rate (**AN-FOR**): probability of a patient having breast cancer if ruled out and the radiologist would have caught it (i.e. N-FOR), adjusted for breast cancer cases that AI retained but radiologists would have missed.

AI False discovery rate (**AI-FDR**): probability of a patient not having breast cancer if retained. AI-FDR = (1-PPV) = FP/ (TP + FP).

Note, TP (true positive), TN (true negative), FP (false positive), FN (false negative).

### Modeling Assumptions

To model AN-FOR, we simulated four hypothetical scenarios in which 10%, 30%, 50%, or 70% of missed cancers in standard practice were detected by using AI triage.

### Statistics

All modeling was conducted using SAS 9.4 (SAS Cary, NC), where sensitivities and specificities were estimated using the %ROCPLOT macro, and PPV, NPV, FDR, and FOR were calculated using Bayes’ Theorem. The base rate of cancer was 0.76%.

## Results

### Approaches to Identifying Triage Threshold

Data were simulated using two triage thresholds that can be generalized across practices. The first uses diagnostic performance—Youden’s J—to define a threshold by optimizing the balance of sensitivity and specificity. The second defines a threshold using an outcome, in this case, avoiding any overall increase in missed breast cancers compared to standard practice without triage. That is, this threshold is set so that an AN-FOR of 0 is achieved, meaning all cancers missed by using AI triage (rule-out cases) is then offset by an identical number of cancer cases that a radiologist would catch because they were retained.

### Identifying Threshold using Diagnostic Performance (Youden’s J)

For these data, we observed that the Youden’s J value is a Mirai score of 0.20, achieving a sensitivity of 74% and a specificity of 75% (see Table 2 and 4). Given a local prevalence of 0.76%, this translated into ruling out 85,220 cases and retaining 29,009 cases, resulting in a CRR of 75% (85,220/114,229). Of these ruled-out cases, 223 had breast cancer and 84,997 did not, thus achieving a G-FOR of 223/85,220 (0.26%). Of the retained cases, 641 had breast cancer and 28,368 did not, thus achieving an AI-FDR of 97.8%.

**Table 2.**
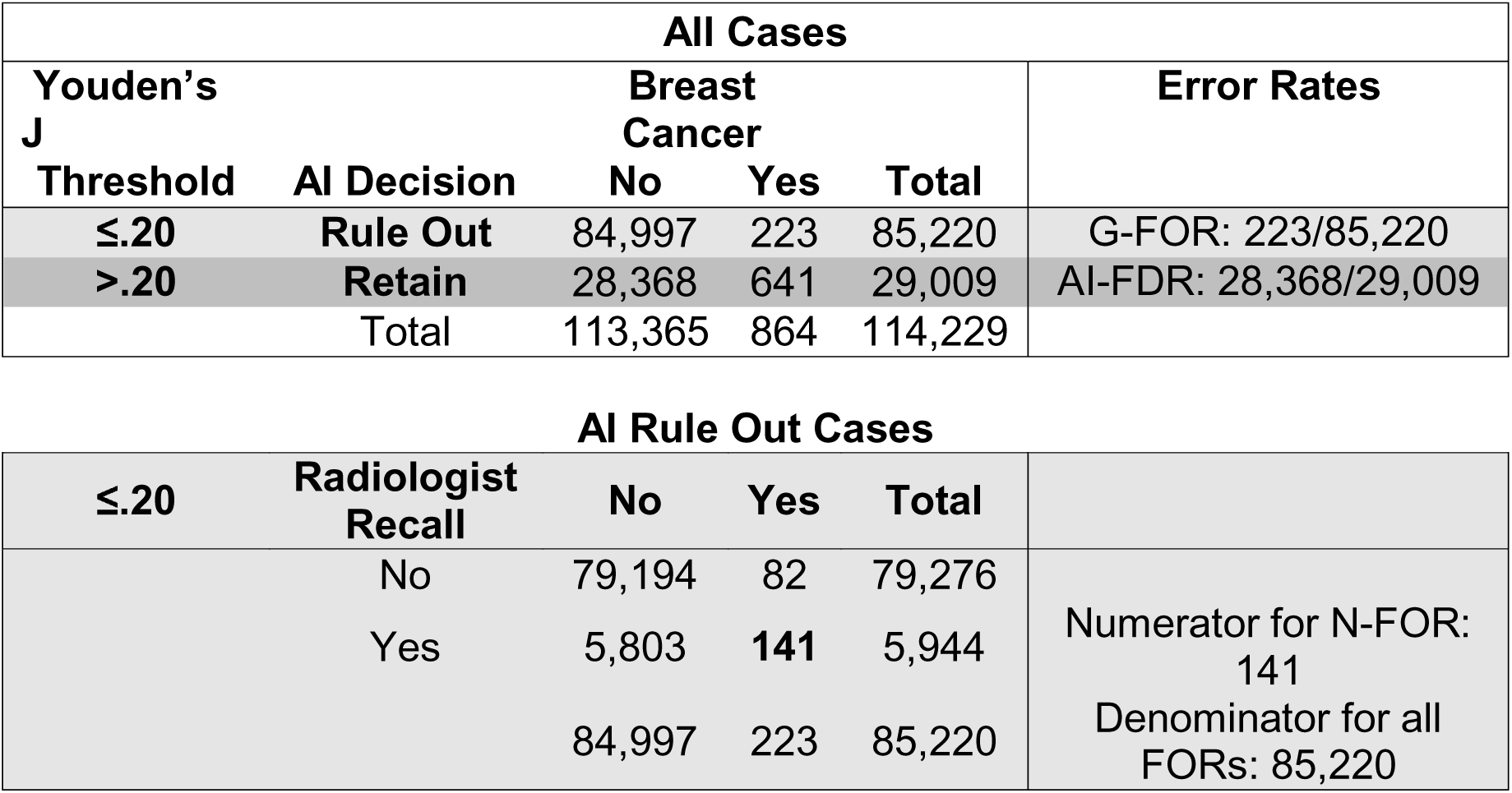
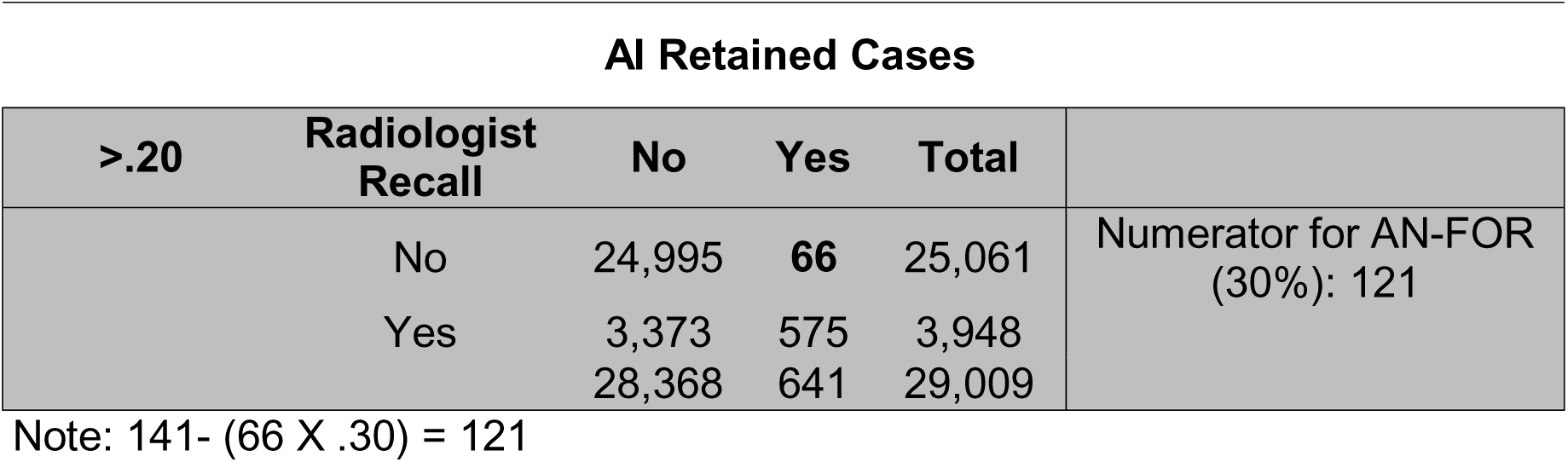
Error Rates Using Youden’s J Threshold.

| All Cases |  |  |  |  |  |
| --- | --- | --- | --- | --- | --- |
| Youden's J Threshold | AI Decision | Breast Cancer |  |  | Error Rates |
|  |  | No | Yes | Total |  |
| ≤.20 | Rule Out | 84,997 | 223 | 85,220 | G-FOR: 223/85,220 |
| >.20 | Retain | 28,368 | 641 | 29,009 | AI-FDR: 28,368/29,009 |
|  | Total | 113,365 | 864 | 114,229 |  |

| AI Rule Out Cases |  |  |  |  |  |
| --- | --- | --- | --- | --- | --- |
| ≤.20 | Radiologist Recall | No | Yes | Total |  |
|  | No | 79,194 | 82 | 79,276 | Numerator for N-FOR: 141<br>Denominator for all FORs: 85,220 |
|  | Yes | 5,803 | 141 | 5,944 |  |
|  |  | 84,997 | 223 | 85,220 |  |

AI Retained Cases
| >.20 | Radiologist Recall | No | Yes | Total |  |
| --- | --- | --- | --- | --- | --- |
|  | No | 24,995 | **66** | 25,061 | Numerator for AN-FOR (30%): 121 |
|  | Yes | 3,373 | 575 | 3,948 |  |
|  |  | 28,368 | 641 | 29,009 |  |

Note: $141 - (66 \times .30) = 121$

Of the 223 breast cancer cases that were ruled out, 82 were not recalled. That is, 82 were also missed by radiologists in standard of care while they recalled the remaining 141, thus achieving an N-FOR of 141/85,220 or 0.17%.

Regarding the retained cases, AI retained 66 cases that radiologists missed. Assuming radiologists detect 10%, 30%, 50% or 70% of these cases in AI triage, the adjusted net number of missed cancers in AI triage would be reduced to by 7, 20, 33, or 46 to 134, 121, 108, or 95, respectively. This would correspond to Adjusted Net FOR values of 0.16%, 0.14%, 0.13%, and 0.11%, respectively. These values are visualized in Table 4 and Figure 1.

**Figure 1.**
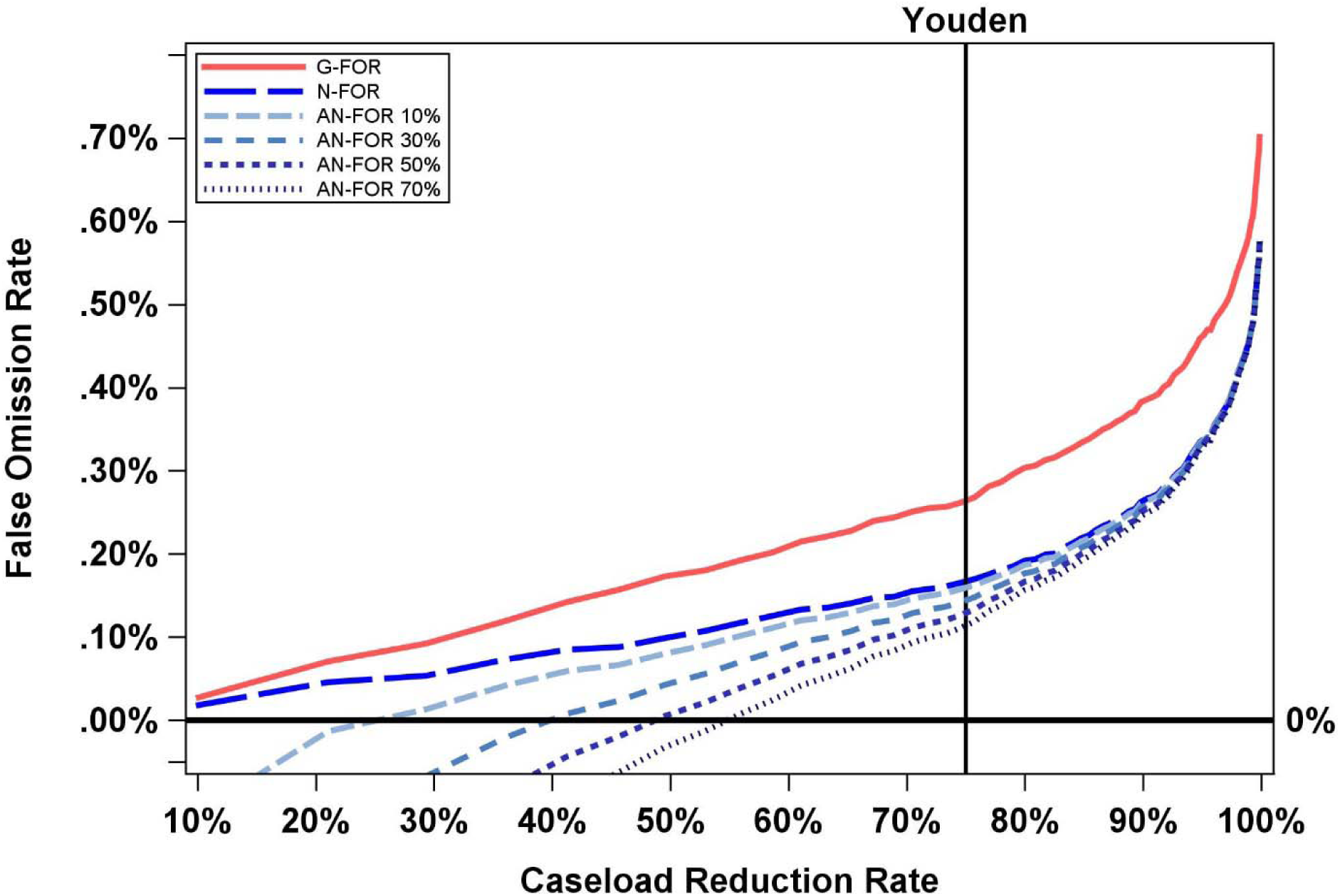
False Omission Rate by Caseload Reduction Rate. X-axis is caseload reduction rate (10% to 100%) and Y-axis is False Omission Rate (0.0% to 0.70%). Youden refers to Youden’s J (thin black line). G-FOR is Gross False Omission Rate (solid red). N-FOR is Net False Omission Rate (longest dash, bright blue). AN-FOR 10% (long dash, light blue), AN-FOR 30% (short dash, medium blue), AN-FOR 50% (short dash, dark blue), AN-FOR 70% (shortest dash, grey blue) refer to the Adjusted Net False Omission Rate at various percentages of additional breast cancers that radiologists would detect (10%, 30%, 50%, and 70% respectively) in AI-retained cases using an AI triage model relative to standard of care.

### Identifying Threshold using Outcomes

Another approach to identifying the threshold is by considering the type of error and number of errors that would result from AI triage based on historical data. As shown in Figure 1 and Tables 3 and 4, depending on the percentage of additional breast cancer cases (i.e., 10%, 30%, 50%, and 70%) that radiologists would have detected among those retained by AI triage (compared to standard of care), the rule-out threshold can be set by determining the caseload reduction rate where AN-FOR intersects a certain value (here 0). As discussed above, this threshold corresponds to no additional missed cancers overall (among both retained and ruled out cases) relative to standard practice. As illustrated in Figure 1 and Table 4 (**bold**), assuming radiologists detect an additional 30% of missed cancers in cases retained by AI, a threshold of Mirai = 0.05 would achieve an AN-FOR of 0, which would translate into a CRR of about 36%. If radiologists detect an additional 70% of missed cancers, a threshold of Mirai=0.09 would achieve an AN-FOR of 0, which would translate into a CRR of about 53%.

**Table 3.**
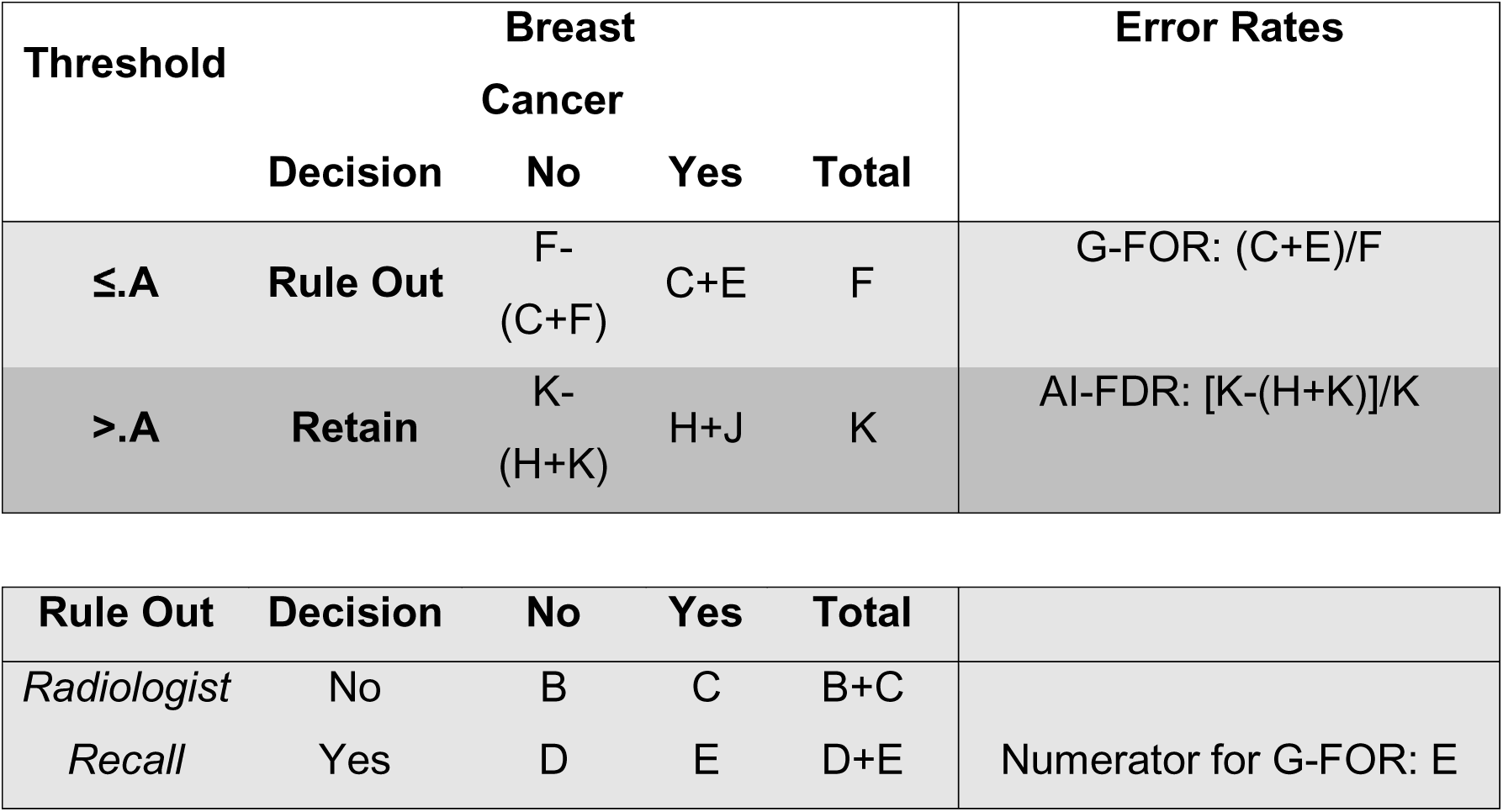
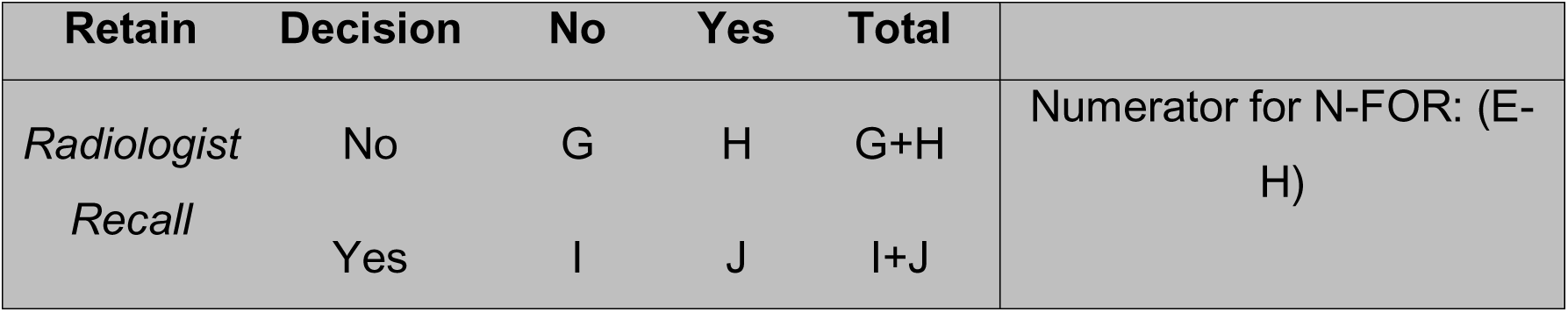
Key for Table 4.

**Table 4.**
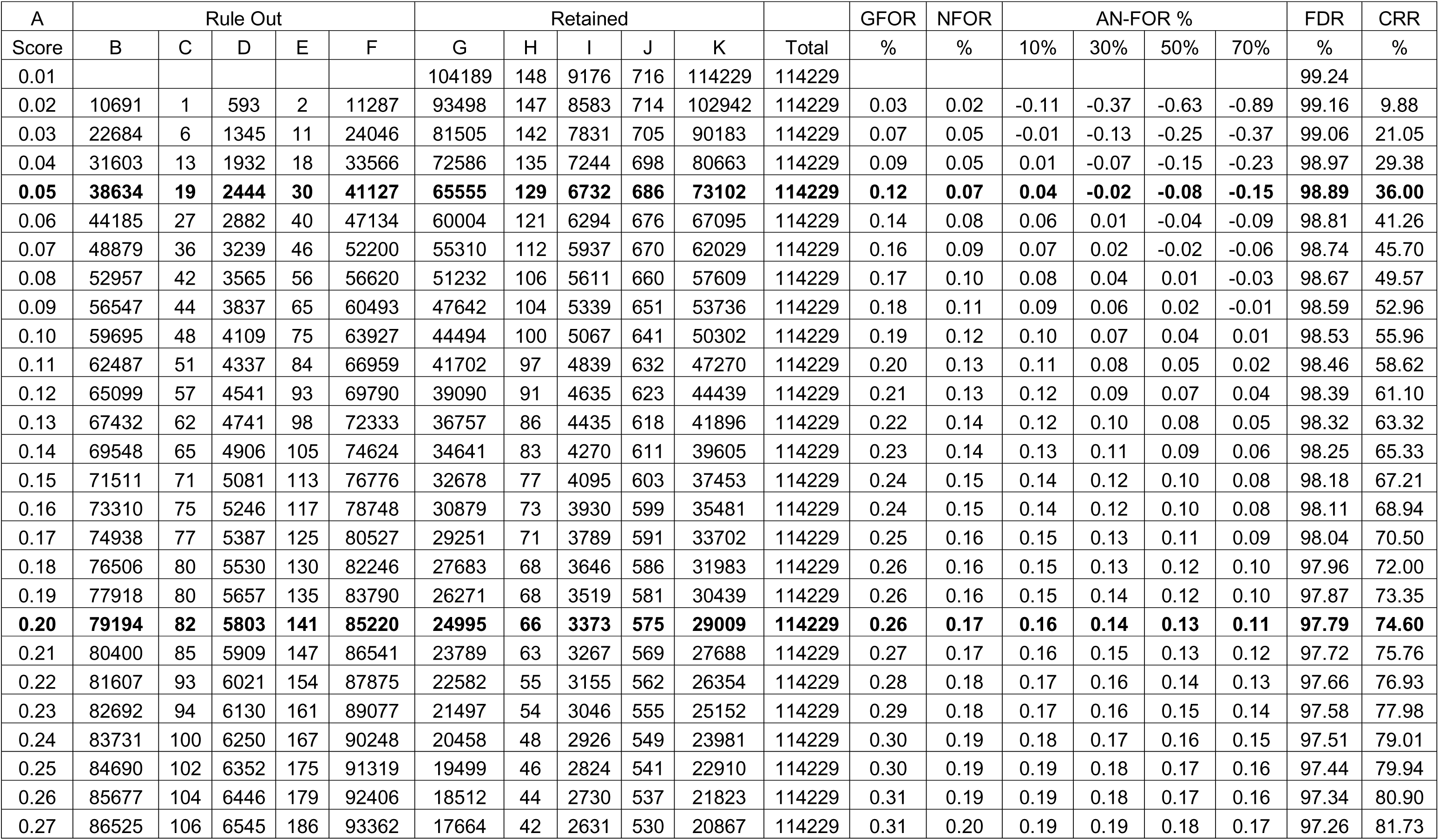
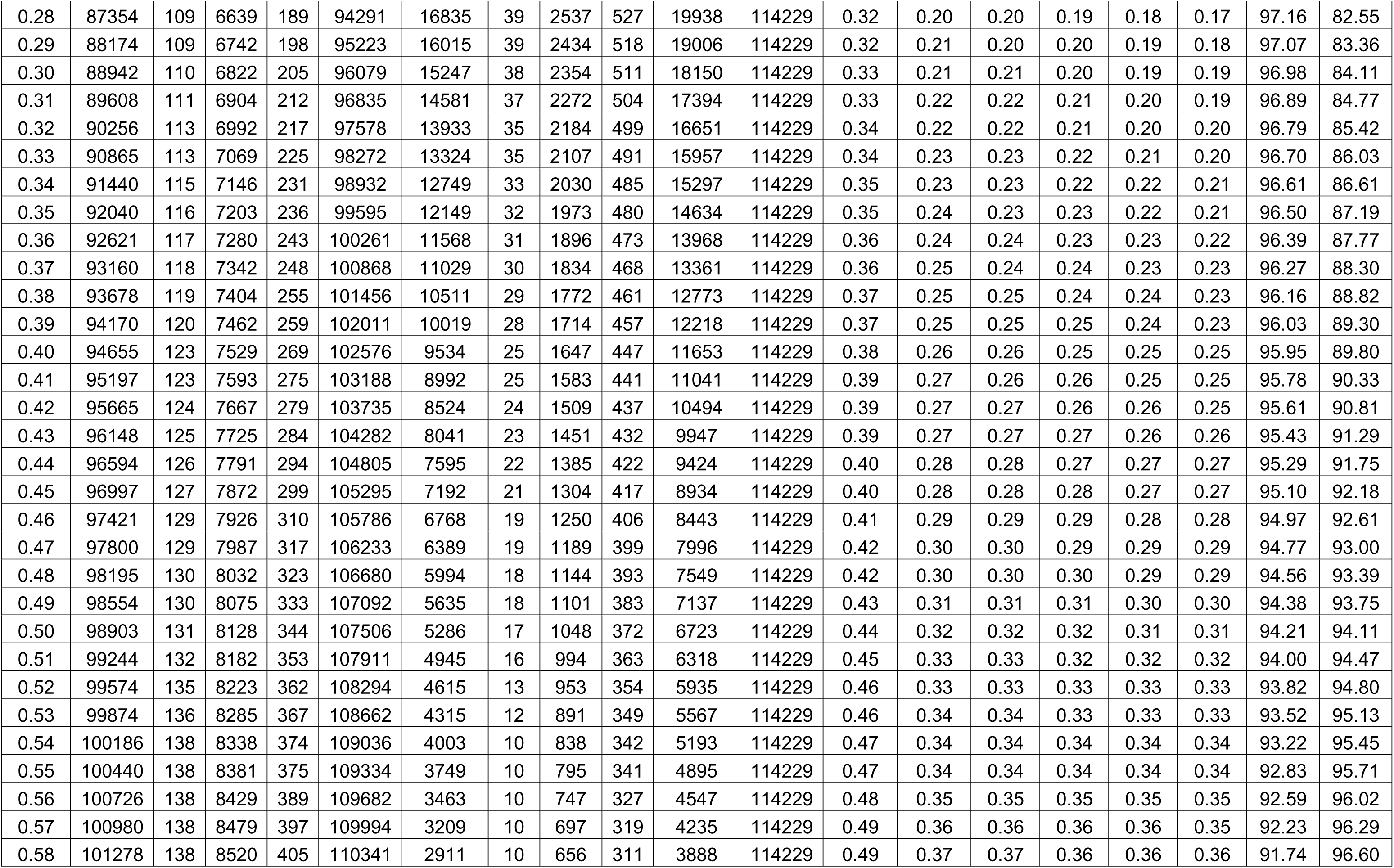
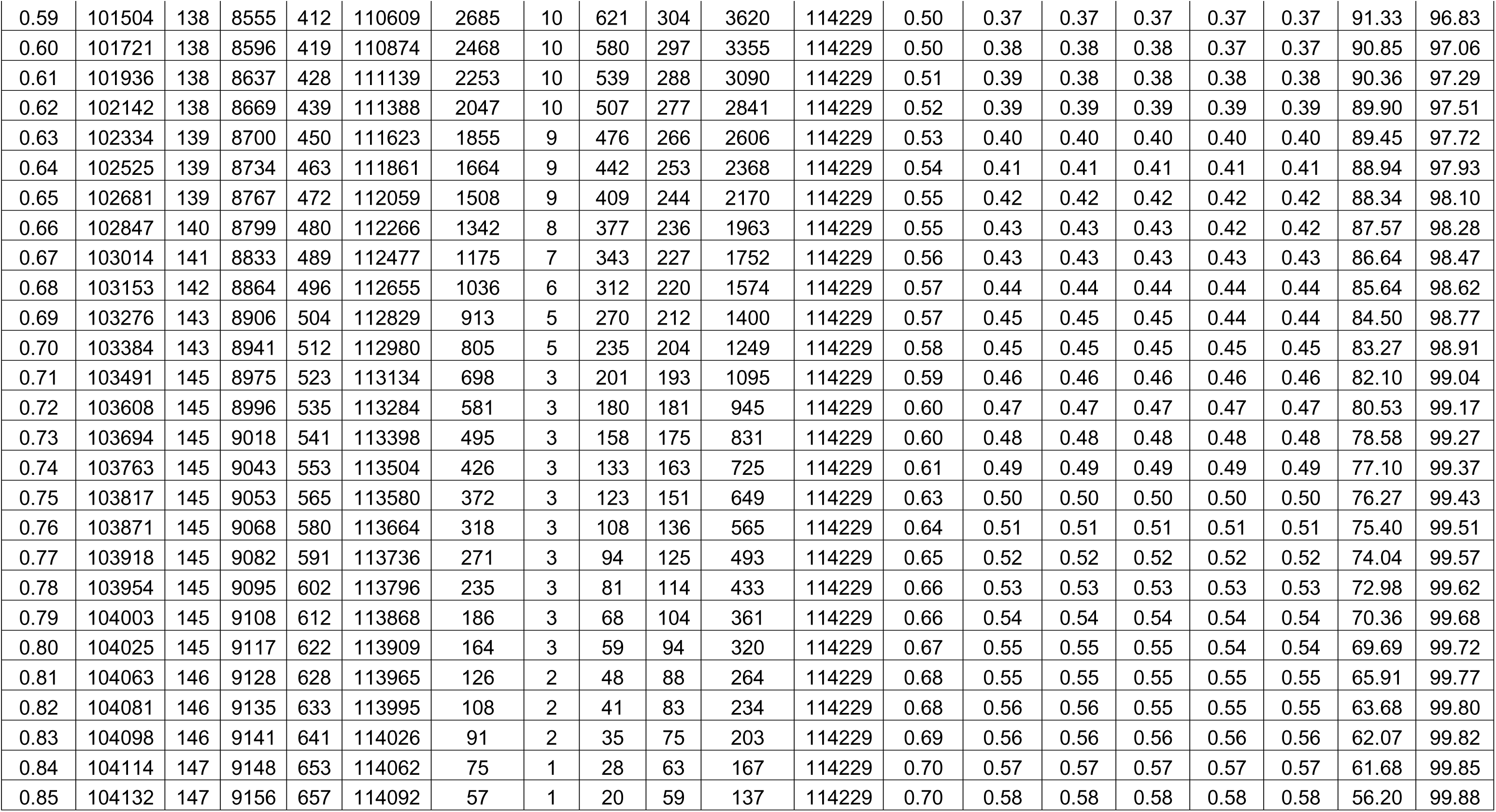
Table of metrics and outcomes.

These CRR values can then be used to examine the corresponding number of false positives. As seen in Figure 2, the AI-FDR was between about 98% and 99% for all thresholds considered, indicating that FDR was largely stable. Given that false positives are unlikely to vary significantly, mainly because of low cancer prevalence,^24^ false negatives will be the primary focus here.

**Figure 2.**
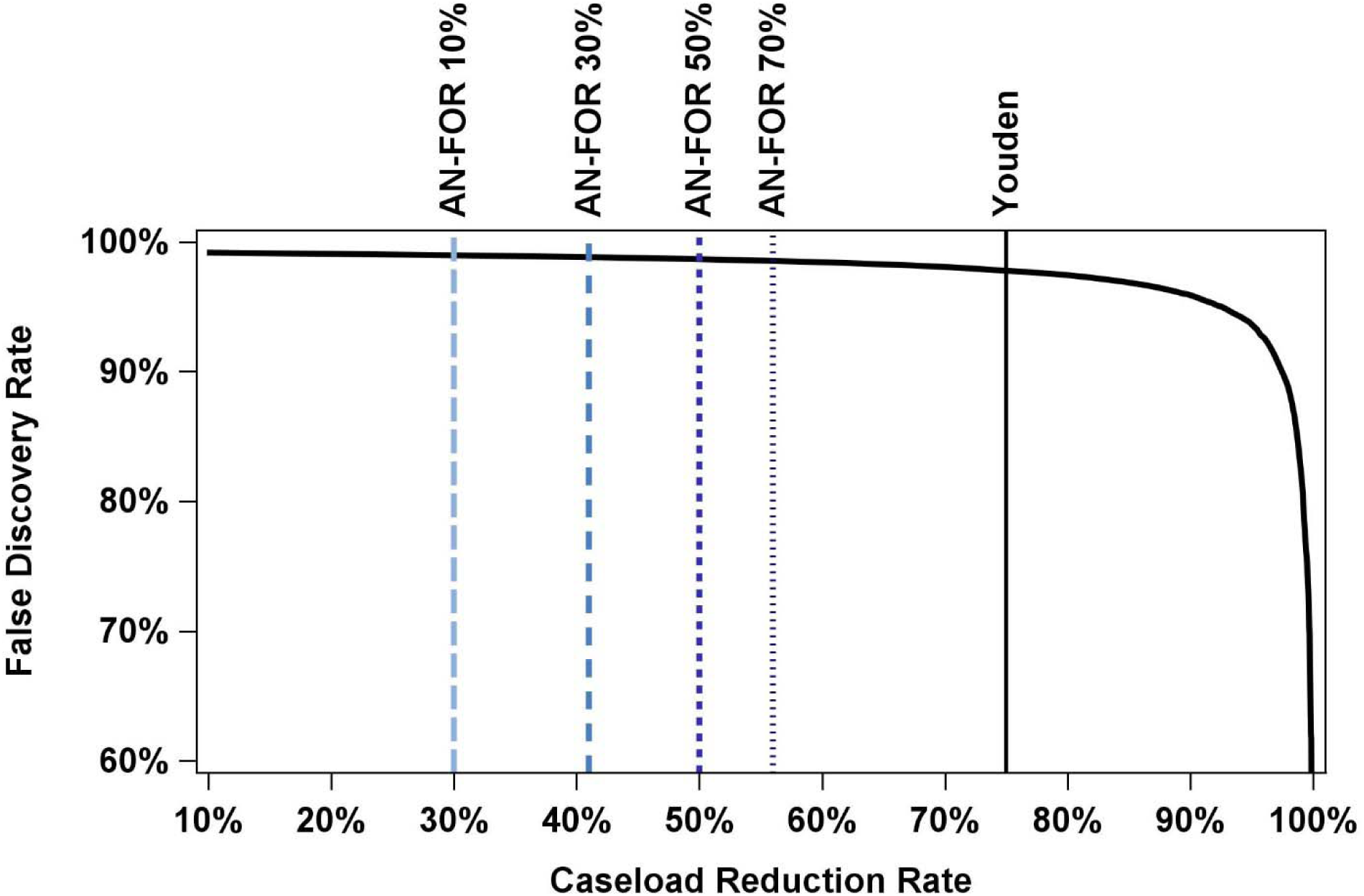
False Discovery Rate by Caseload Reduction Rate. X-axis is caseload reduction rate (10% to 100%) and Y-axis is False Discovery Rate (60% to 100%). Thick black line is the relationship between Caseload Reduction Rate and False Discovery Rate Youden refers to Youden’s J (thin black line). AN-FOR 10% (long dash, light blue), AN-FOR 30% (short dash, medium blue), AN-FOR 50% (short dash, dark blue), AN-FOR 70% (shortest dash, grey blue) refer to the Adjusted Net False Omission Rate at various percentages of additional breast cancers that radiologists would detect (10%, 30%, 50%, and 70% respectively) in AI-retained cases using an AI triage model relative to standard of care.

### Comparing Thresholds

To assess the trade-off between errors and benefits, we compare two thresholds: Mirai score of 0.20 corresponding to a 75% CRR (Youden’s J) and Mirai score of 0.05 corresponding to a 36% caseload reduction assuming AN-FOR of 0 where 30% of additional breast cancers would have been detected among retained cases by radiologists using an AI triage model compared to standard of care.

At the 0.20 threshold, the G-FOR, N-FOR, and AN-FOR (30%) are 0.26%, 0.017%, and 0.14%, respectively. This corresponds to 223, 141, and 121 missed cancer cases for the benefit of reading 85,220 fewer cases with an FDR of 97.8%. In contrast, at the 0.05 threshold, the G-FOR, N-FOR, and AN-FOR (30%) are 0.12%, 0.07%, and 0.00% (rounded), corresponding to 49, 30, and 0 missed cancer cases for the benefit of reading 41,127 fewer cases with an FDR of 98.9%. Tables 3 and 4 provide all combinations for comparison.

## Discussion

We demonstrate how radiology practices can consider the trade-offs of using different AI scores to determine the triage threshold. Using the Mirai AI algorithm and historical data, our simulation demonstrates how a risk-benefit analysis could be quantified. The purpose of this framework is not to advocate for a specific threshold or risk-benefit ratio but rather to demonstrate how a risk-benefit ratio could be quantified to inform policy and clinical implementation of AI triage. All numerical values provided are illustrative and are not intended as recommendations for clinical use.

The optimal threshold will vary depending on the AI model, the pathology (and the corresponding trade-offs of false positives and false negatives), the AI model’s sensitivity and specificity for a local population, the prevalence of the local population, the local caseload volume and radiologist staffing ability, and institutional risk tolerances. Our simulation highlights how error rates (risk) and caseload reduction rate (benefit) can be estimated using historical data. This estimation not only accounts for the **type of errors (i.e., false positive and false negative)** but also the **number of errors (i.e., false discovery and omission rates instead of false positive and negative rates)**.

While our simulation focused on the number of any missed cancers, the type (e.g. in situ versus invasive) and stages of cancers missed by AI could be incorporated to further assess the clinical significance of triage-related errors. What is more, we only evaluated cancers diagnosed within a year of the screening mammogram; other time frames (e.g., 1 and 2-year cancer outcomes) could be incorporated as well. Finally, for simplicity, we calculated the G-FOR, N-FOR, AN-FOR, and FDR using the direct rates, although confidence, prediction, or credible interval estimates could be used instead. Again, the point of the current study is to demonstrate the general framework of how triage could be used relative to standard of care.

Fan et al. also propose evaluating AI triage using PPV and NPV. Our approach builds upon Fan and colleagues in two key ways. Namely, Fan et al. do not account for key counterfactuals such as cancers that would have been missed by radiologists without triage and cancer only detected with triage because of changes in radiologist performance.^16^ In addition, Fan et al. propose using expected utility (EU) to assess AI triage. However, this relies on baseline relative utility values, which are difficult to define and when defined, may be difficult to justify, economically, ethically, and otherwise.

Along with a framework for determining a threshold for AI triage of screening mammograms, there are several important considerations that must be addressed before AI triage can be implemented in clinical practice. First, prospective validation of AI rule-out strategies is needed. This validation will be important for understanding how AI triage impacts radiologist performance in the retained cases, such as increased recalls and increased cancer detection to empirically determine AN-FOR (whereas we simulated potential values). The validation will also be critical for ensuring that AI triage performs equitably across patient groups. Second, standards need to be developed for the safe deployment of AI triage tools in clinical settings and address approaches for ongoing monitoring of AI performance and safety over time. Third, there are psychological, ethical, legal, economic, and insurance considerations that must be weighed if implementing triage. Finally, there will need to be significant changes to the policy and regulatory landscape to allow AI triage in clinical practice. Addressing these considerations is necessary for the implementation of AI triage.

## Conclusion

We present a framework for quantifying AI triage thresholds based on errors and benefits. Such a framework can help translate the potential of AI into strategies that help alleviate the growing workload pressures and resource limitations in radiology.

## Data Availability

All data produced in the present study are available upon reasonable request to the authors

